# Stakeholder perspectives and preferences for long-acting injectables for tuberculosis treatment and prevention: a multi-regional mixed-methods study

**DOI:** 10.64898/2026.11.06.26367430

**Authors:** Chaelin Kim, Erin V. McConnell, Samuel G. Schumacher, Dennis Falzon, Medea Gegia, Lusiana Aprilawati, Jimmy Galarza, Edna Tembo, Lesia Tonkonoh, Gwenan M Knight, C Finn McQuaid, Fuad Mirzayev, Nora West

**Author notes:** Joint first authors (Chaelin Kim, Erin V. McConnell). Joint senior authors (Fuad Mirzayev, Nora West).

## Abstract

**Objective:** To assess stakeholder acceptability of and preferences for long-acting injectables (LAIs) for tuberculosis (TB) treatment and prevention across World Health Organization regions to inform product development and implementation.

**Methods:** We conducted a sequential explanatory mixed-methods study of survey data and qualitative focus group discussions (FGDs). Online surveys assessed perceptions and acceptance of LAIs among two groups: TB programme managers, civil society members and healthcare workers (Survey A), and community members and persons affected by TB (Survey B). Using purposive sampling, survey participants were invited to join FGDs to explore survey findings. Survey data were analysed using descriptive statistics, ordinal logistic regression with stepwise AIC selection, and one-sample proportion tests. FGD transcripts were analysed using modified framework analysis.

**Findings:** Among 310 participants (102 Survey A; 208 Survey B), 83% (n=256) favoured LAIs over standard oral TB treatment. Compared with six months of daily oral treatment, 69% preferred a three-month regimen comprising one month of daily oral treatment followed by four injections per monthly visit for two months, while 80% preferred the corresponding regimen with two injections per visit. For prevention, 79% preferred a single injection over one month of oral treatment. In Survey B, interest varied according to lived TB experience, with FGDs suggesting that previous negative experiences with injectables may have contributed to this variation.

**Conclusion:** Interest in LAIs for TB treatment and prevention was high, but lower among participants with lived TB experience. Product development and implementation should address concerns about injectables and side effects while preserving patient choice and agency.

## Introduction

Tuberculosis (TB) remains a leading cause of infectious disease morbidity and mortality worldwide (1). Although treatment regimens have become shorter and safer, both treatment of disease and TB preventive treatment (TPT) continue to rely largely on the intake of multiple oral medicines taken daily over several months. Adherence to these regimens is influenced by the long treatment duration and pill burden as well as side effects, stigma, competing personal responsibilities, transportation costs, health literacy, social support, and health service accessibility and quality (2–4). Poor initiation or interruption of medication can reduce the effectiveness of TB treatment and contribute to prolonged infectiousness, unfavourable outcomes, relapse, and the emergence of drug resistance (5,6).

Long-acting injectables (LAIs) could offer an alternative to taking pills daily. By reducing the frequency of medicine administration, LAIs may decrease pill burden, treatment fatigue, and the time and resources required to manage repeated dosing, leading to increased uptake and treatment success (7,8). Experience with existing long-acting formulations for contraception, psychiatry, and HIV management has demonstrated that extended-release delivery can be acceptable, effective, and capable of supporting treatment continuation (9–12). Several long-acting formulations of TB medicines are now in preclinical or early clinical development for TPT and the treatment of TB disease (8,13–18).

The acceptability and programmatic value of an LAI regimen will depend on more than its pharmacological efficacy. The number and frequency of injections, injection volume, route and anatomical site of administration, treatment duration, location of care, and the inclusion of oral medicines create trade-offs for patients. For TB treatment, which requires combination therapy over several months, the ideal LAI regimen would deliver all requisite medicines in a single injection. However, the co-formulation of multiple long-acting agents into a single injection is technically challenging, meaning that TB treatment with LAIs may require multiple injections per visit and/or an unsynchronized treatment and dosing schedule. This creates a central design tension between minimising the number of injections and injection volume, and the practical need to deliver multiple agents for treatment. Fewer dosing encounters may simplify treatment, but large-volume or multiple injections at a single visit may increase pain and injection-site reactions.

Extant literature from India, South Africa, and the Philippines shows favourable attitudes towards LAIs for TPT and TB treatment, with preferences shaped by efficacy and side-effect concerns, injection number, site and pain, and place of delivery (19,20). These findings suggest that acceptability will depend on specific regimen characteristics, making early engagement with intended users essential. However, both studies were limited to one or two countries and to either prevention or treatment, and neither explored how contextual factors shape preferences among people affected by TB. We therefore conducted a sequential explanatory mixed-methods study involving stakeholders from multiple World Health Organization (WHO) regions to examine perceptions of LAIs for TB treatment and prevention and preferences for key regimen characteristics.

## Methods

### Study Design

This study employed a sequential explanatory mixed-methods design among adults 18 years or older, comprising an initial quantitative phase followed by a qualitative phase designed to explore and contextualize the survey findings. Phase 1 consisted of online cross-sectional surveys assessing preferences for and acceptability of LAIs for TB prevention and treatment across all six WHO regions. Following survey data collection and initial analysis, Phase 2 comprised four regionally organized focus group discussions (FGDs). Quantitative findings were used to inform the FGD facilitation guide, with discussions focusing on areas of consensus, heterogeneity in preferences and concerns across geographic and sociocultural contexts, and unexpected findings requiring further exploration. Integration occurred at the interpretation stage, with qualitative findings used to explain and contextualize quantitative patterns in a joint narrative synthesis. Further details on survey instrument development, qualitative data collection, analytic rigor and reflexivity, and mixed-methods integration are provided in **Appendix A**.

### Ethics

This study received ethical approval from the WCG Institutional Review Board (IRB Protocol #: 20252075) and the London School of Hygiene & Tropical Medicine Research Ethics Committee (LSHTM Ethics Ref: 33795).

### Phase 1: Quantitative Survey

#### Sampling and Recruitment

The Phase 1 surveys used complementary sampling methods to gather perspectives from TB service delivery stakeholders and TB-affected communities. Survey A targeted healthcare, policy, and civil society stakeholders, while Survey B targeted TB-affected communities, including individuals with lived TB experience. Recruitment was conducted via email and listservs. To increase geographic diversity, purposive snowball sampling was used through regional TB networks and direct outreach to community representatives working with marginalized and hard-to-reach populations in South America, Francophone Africa, Eastern Europe, and Central Asia. The surveys were administered through Google Forms and collected anonymous, voluntary responses without any personally identifying information.

#### Quantitative Analysis

Survey data were analysed using descriptive statistics, one-sample proportion tests, and ordinal logistic regression. Participant characteristics and LAI interest responses were summarized using frequencies, percentages, means, and standard deviations, and were stratified by stakeholder affiliation, WHO geographic region, and prior TB-related experience. Interest in LAIs for TB treatment was measured on a five-point ordered response scale: 1 (“Definitely not”), 2 (“Probably not”), 3 (“I am unsure”), 4 (“Probably”), and 5 (“Definitely”). For items where Survey A (Provider/Policy Survey) and Survey B (Community/Patient Survey) used identical questions and response options, data were combined to create a larger, more stakeholder-diverse dataset (total N = 310).

Factors associated with interest in LAIs for TB treatment were analyzed using ordinal logistic regression, with the five-level LAI interest variable treated as an ordered outcome. Univariate analyses were conducted for each predictor, and crude odds ratios (CORs), 95% confidence intervals (CIs), and p-values were estimated. We selected variables for the multivariable model using stepwise Akaike Information Criterion (AIC)-based selection, including backward, forward, and bidirectional approaches. The final model included variables that minimized the AIC (**Appendix B**). Adjusted odds ratios (AORs), 95% CIs, and p-values were reported for variables retained in the final multivariable model. For regression analysis of factors associated with LAI interest, only Survey B data were used (n = 205) because this survey included additional variables on participant characteristics not captured in Survey A.

Treatment preference between paired regimens was assessed using one-sample proportion tests, comparing the observed proportion favoring the preferred option to the null hypothesis of 50%. For these analyses, responses were categorized as Prefer Option A, Neutral, or Prefer Option B by combining strong and moderate preferences for each option. The preferred option was defined as the option with the highest combined preference, and proportions were reported with 95% CIs. All quantitative analyses were conducted in R.

### Phase 2: Qualitative Focus Group Discussions

#### Sampling and Recruitment

Eligibility criteria for the FGDs required participants to have previously completed the Phase 1 community survey and to demonstrate proficiency in English (focus groups were conducted in English). We recruited participants via email outreach to individuals who indicated in Phase 1 that they were willing to be contacted for follow-up participation. To ensure geographic representation, targeted purposive outreach was used in regions with a smaller pool of eligible participants, including Eastern Europe and Central Asia, and the Region of the Americas.

Participants were organized into four regionally defined focus groups reflecting WHO regional groupings: African Region (AFRO); Region of the Americas (AMRO); Western Pacific and South-East Asia Regions (WPRO & SEARO); and European and Eastern Mediterranean Regions (EURO & EMRO). Each group targeted up to ten participants (total target N = 40). Participants provided consent via Google Forms prior to attendance. Semi-structured FGDs were conducted online and recorded with participant consent.

#### Qualitative Analysis

Focus group transcripts were transcribed by N.S.W and were analyzed using a modified framework analysis approach (Gale et al., 2013). Following transcription, transcripts were deidentified and imported into ATLAS.ti software (21) for coding. Analytical coding was organized within three a priori domains derived from the study’s objectives and the Phase 1 survey structure: (1) LAI acceptability, (2) LAI treatment characteristic preferences, and (3) community-specific considerations for implementation. Within these domains and based on data familiarization, codes and emergent themes were further developed and discussed among the study team (N.S.W. and E.V.M.).

All transcripts were coded and charted into an analytic matrix by N.S.W.. Transcripts were first summarized by regional groups to identify within-region patterns, and findings were subsequently charted across regions to identify convergent and divergent themes. Where regional differences were substantively meaningful, these are noted in the results. Representative participant quotes are presented alongside narrative text to illustrate key themes and preserve the voices of community participants. All participants were assigned pseudonyms for anonymity.

### Mixed Methods Integration

Integration of the quantitative and qualitative data occurred at the interpretation stage, consistent with the explanatory sequential design. Quantitative survey findings established the prevalence and distribution of preferences, acceptability levels, and areas of variation across stakeholder and regional subgroups. Qualitative FGD findings were then used to provide explanatory depth, illuminating the mechanisms, values, lived experiences, and contextual conditions that account for the quantitative patterns. In the results, quantitative data are presented first, followed by qualitative findings, with explicit connections drawn between the two data sources in the mixed methods findings section.

## Results

### Demographics

A total of 310 participants completed the Phase 1 surveys, with 102 participants in Survey A and 208 in Survey B. These participants represented 55 countries across all six WHO regions, with regional representation varying between the two surveys, reflecting their complementary recruitment strategies and target populations (**Table 1**). Fourteen Survey B respondents who agreed to follow-up participated in the FGDs. Further participant characteristics are provided in **Appendix C**.

**Table 1.** Participant demographics by WHO region and qualitative focus group subset.

| **WHO region / FGD region\*** | **Survey A<br>(N=102)** | **Survey B<br>(N=208)** | **Total survey sample<br>(N=310)** | **Qualitative FGD subset<br>(N=14)** |
|---|---|---|---|---|
| Africa | 11 (11%) | 91 (44%) | 102 (33%) | 4 |
| Americas | 3 (3%) | 30 (14%) | 33 (11%) | 3 |
| Europe & Eastern Mediterranean | 27 (26%) / 0 (0%) | 49 (24%) / 2 (1%) | 76 (25%) / 2 (1%) | 2 |
| South-East Asia & Western Pacific | 17 (17%) / 34 (33%) | 30 (14%) / 6 (3%) | 47 (15%) / 40 (13%) | 5 |
| Global | 10 (10%) | 0 (0%) | 10 (3%) | — |

***Note:*** *Qualitative FGD participants were recruited from Survey B respondents who indicated willingness to be contacted for follow-up participation; they therefore represent a subset of the community survey sample. Percentages may not sum to 100% due to rounding. FGD = focus group discussion; WHO = World Health Organization.*
*\* Combined rows reflect the regional categories used for focus group discussions (purposive sampling); survey figures are shown separately for each WHO region.*

### Phase 1 results

Overall interest in LAIs for TB treatment was high across the combined survey sample. Of 310 respondents, 256 (82.6%) indicated probable or definite interest in LAIs for TB treatment, 25 (8.1%) reported that they would probably or definitely not be interested, and 29 (9.4%) were unsure. The mean interest score was 4.16 (SD 0.98) on a five-point scale, where a score of 5 indicated “Definitely”. Descriptive results showed generally high interest across stakeholder groups and WHO regions (**Figure 1, Appendix D**). Ordinal logistic regression examined factors associated with greater interest in LAIs for TB treatment among community survey respondents with complete covariate data (n=205). In the final multivariable model, selected using stepwise AIC, respondents with lived experience of TB had lower odds of reporting greater interest in LAIs compared to those without such experience (AOR 0.51, 95% CI 0.29–0.91) (**Appendix E**).

**Figure 1.**
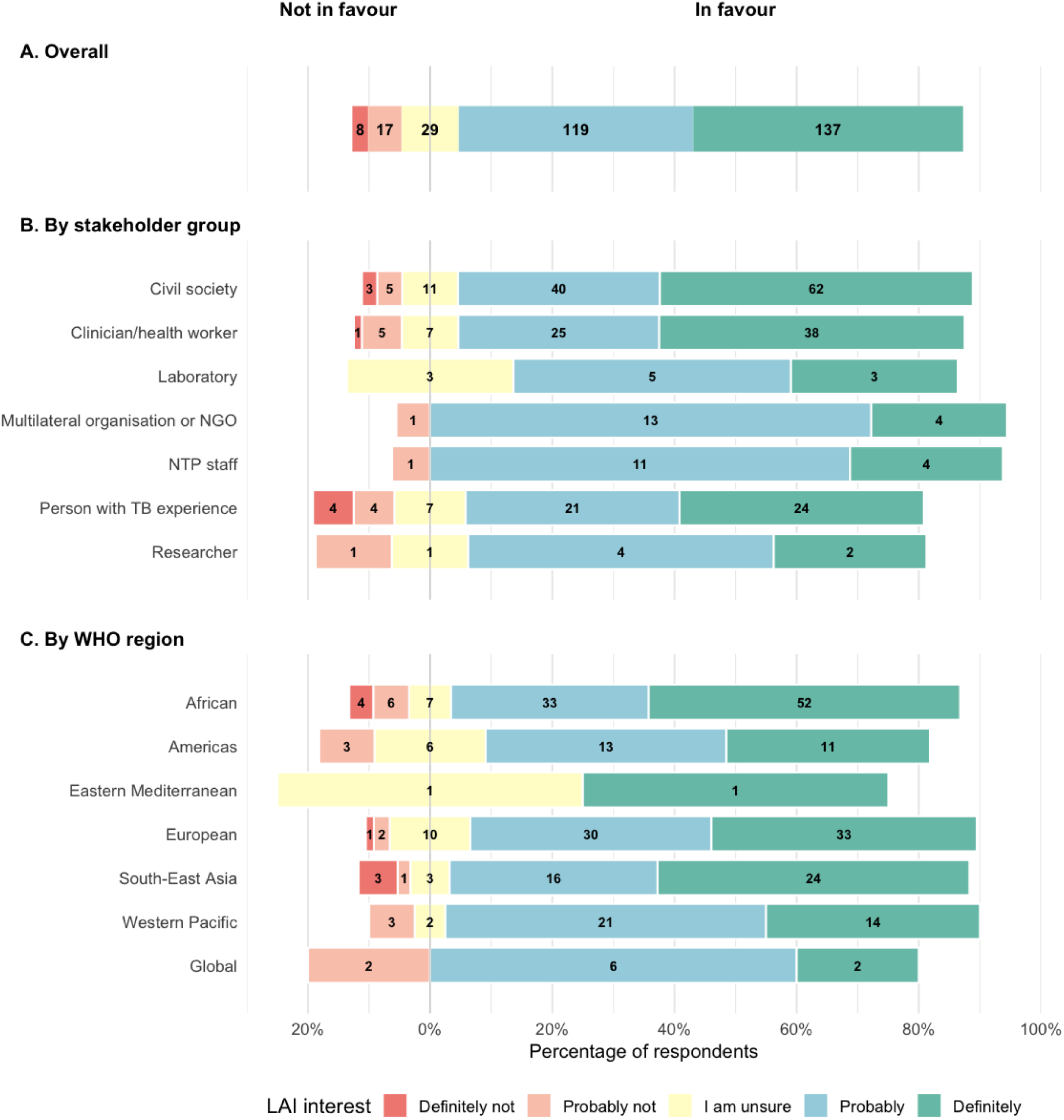
Interest in long-acting injectable regimens for tuberculosis treatment, overall and by stakeholder group and WHO region.

In paired treatment preference comparisons, respondents generally favored LAI regimens when these reduced treatment duration or injection burden. Compared with a 6-month daily oral regimen, 69.4% preferred a 3-month LAI-containing regimen with four injections per month, increasing to 80.3% when injection frequency was reduced to two per month (both p<0.001) (**Table 2**). For TB preventive treatment, 79.4% preferred a single injection over a 1-month oral regimen (p<0.001). Preferences were less clear when options involved competing trade-offs in treatment duration, injection number, route, or location of care. However, 63.9% preferred a shorter intramuscular regimen over a longer subcutaneous regimen (p<0.001). As several paired comparisons varied multiple attributes simultaneously, these findings reflect combined regimen preferences rather than isolated effects of individual characteristics (full question wording and illustrations in **Appendix F**).

**Table 2.**
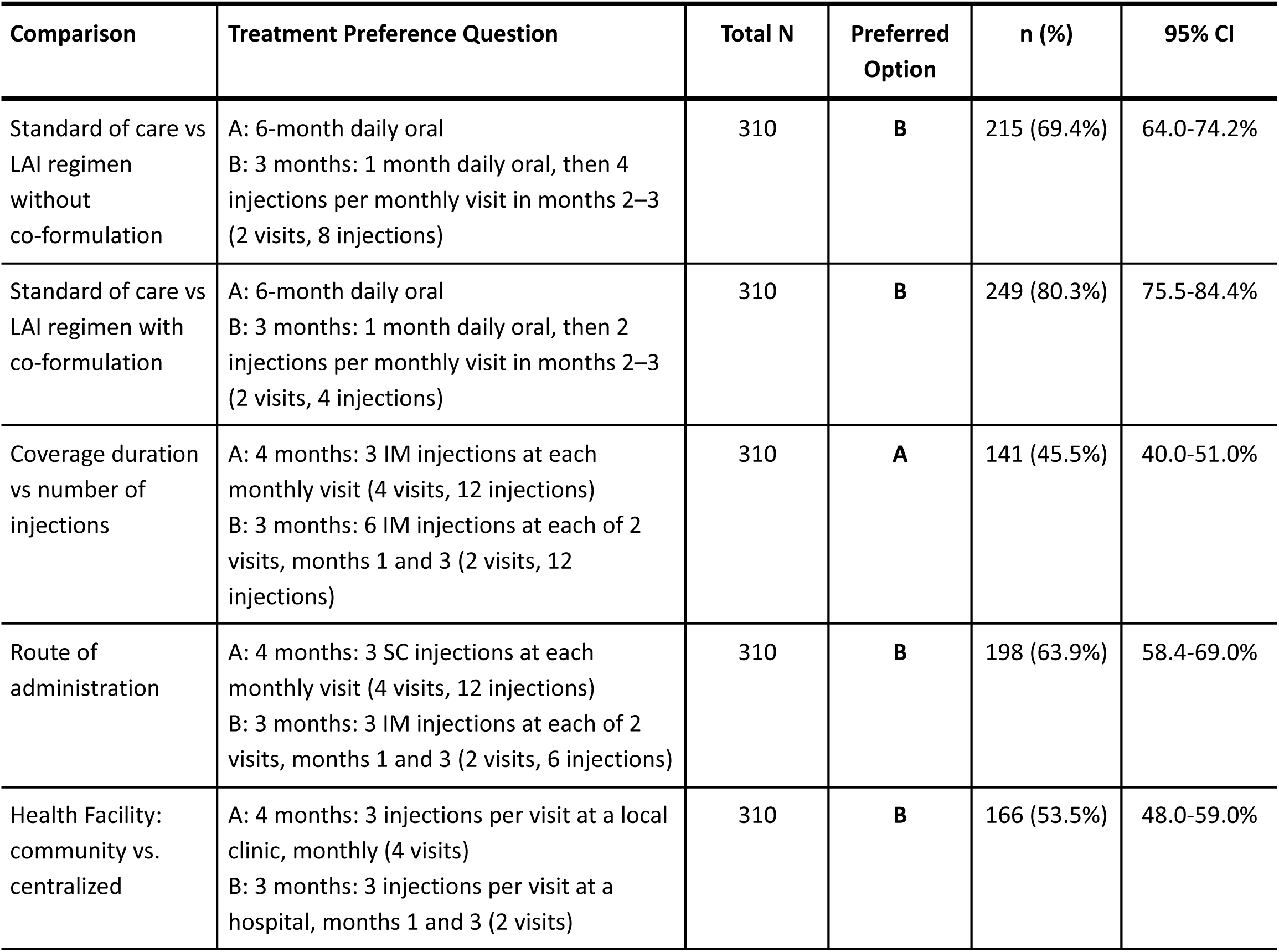
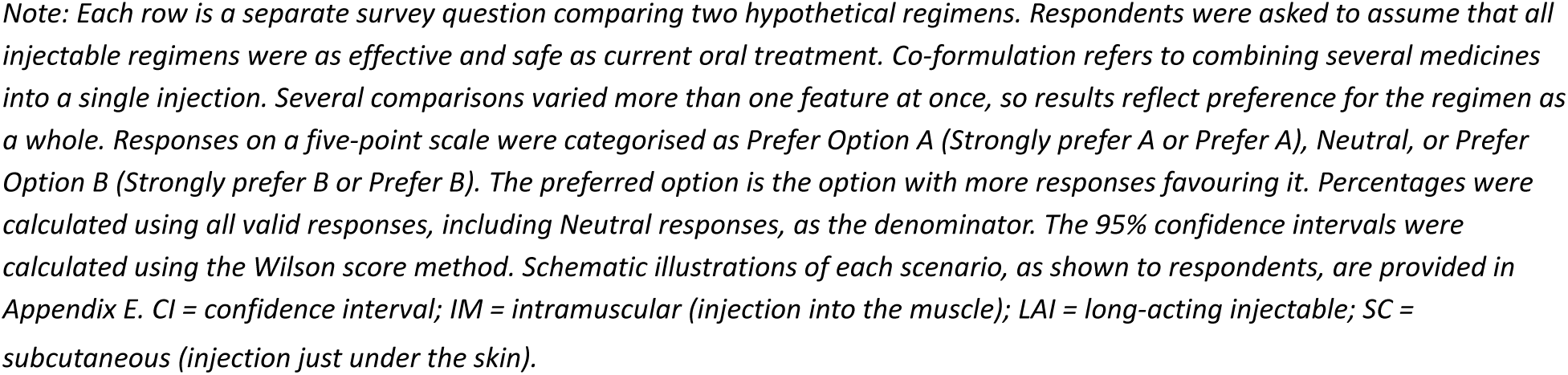
Treatment Preferences for Long-Acting Injectable TB Regimens.

| Comparison | Treatment Preference Question | Total N | Preferred Option | n (%) | 95% CI |
|---|---|---|---|---|---|
| Standard of care vs LAI regimen without co-formulation | A: 6-month daily oral<br>B: 3 months: 1 month daily oral, then 4 injections per monthly visit in months 2–3 (2 visits, 8 injections) | 310 | **B** | 215 (69.4%) | 64.0-74.2% |
| Standard of care vs LAI regimen with co-formulation | A: 6-month daily oral<br>B: 3 months: 1 month daily oral, then 2 injections per monthly visit in months 2–3 (2 visits, 4 injections) | 310 | **B** | 249 (80.3%) | 75.5-84.4% |
| Coverage duration vs number of injections | A: 4 months: 3 IM injections at each monthly visit (4 visits, 12 injections)<br>B: 3 months: 6 IM injections at each of 2 visits, months 1 and 3 (2 visits, 12 injections) | 310 | **A** | 141 (45.5%) | 40.0-51.0% |
| Route of administration | A: 4 months: 3 SC injections at each monthly visit (4 visits, 12 injections)<br>B: 3 months: 3 IM injections at each of 2 visits, months 1 and 3 (2 visits, 6 injections) | 310 | **B** | 198 (63.9%) | 58.4-69.0% |
| Health Facility: community vs. centralized | A: 4 months: 3 injections per visit at a local clinic, monthly (4 visits)<br>B: 3 months: 3 injections per visit at a hospital, months 1 and 3 (2 visits) | 310 | **B** | 166 (53.5%) | 48.0-59.0% |

*Note: Each row is a separate survey question comparing two hypothetical regimens. Respondents were asked to assume that all injectable regimens were as effective and safe as current oral treatment. Co-formulation refers to combining several medicines into a single injection. Several comparisons varied more than one feature at once, so results reflect preference for the regimen as a whole. Responses on a five-point scale were categorised as Prefer Option A (Strongly prefer A or Prefer A), Neutral, or Prefer Option B (Strongly prefer B or Prefer B). The preferred option is the option with more responses favouring it. Percentages were calculated using all valid responses, including Neutral responses, as the denominator. The 95% confidence intervals were calculated using the Wilson score method. Schematic illustrations of each scenario, as shown to respondents, are provided in Appendix E. CI = confidence interval; IM = intramuscular (injection into the muscle); LAI = long-acting injectable; SC = subcutaneous (injection just under the skin).*

### Phase 2 Results

Community perspectives on long-acting injectables for TB were broadly favorable across all four regions (**Table 3**), with acceptability grounded primarily in relief from pill burden and the simplification of treatment regimens. Participants felt that overall fewer injections at longer intervals could address adherence challenges associated with daily oral medications, particularly in contexts where food insecurity or work schedules interfere with pill-taking. However, this acceptance was nuanced: historical memories of severe side effects from previous injectable regimens for MDR-TB continue to influence perceptions. Gender differences also emerged, with participants discussing concerns about modesty related to buttocks injections for men and that women have greater engagement with health systems overall, making LAIs potentially more accessible for them.

**Table 3.** Qualitative themes, key findings, and representative quotes on long-acting injectables for tuberculosis.

| Theme | Key Finding | Representative Quote |
|---|---|---|
| **Domain: Acceptability** |  |  |
| **Overall Acceptability of LAIs** | Broadly favorable across all regions, grounded in relief from pill burden and simplified dosing | *"I think this [LAIs] will be a great milestone because most of the time, people have complained of having to swallow a lot of pills once they have TB. So, if the injections are there, they are receiving them once every three months or every two months...if it is an injection, sometimes people will not directly think of getting food now...because the injection will be acting slowly as compared to the pills, which I cannot swallow on an empty stomach." —African Region Participant* |
| **Historical Injectable Stigma (MDR-TB)** | Legacy of severe side effects from Multi-Drug Resistant (MDR-TB) injectables shapes current perceptions | *"At the same time, if coming back to those times when people with TB didn't have any opportunity to choose while having TB treatment...the injections were painful, and at the same time, they had lots of adverse effects...the new generation...have the experience of communicating with people with TB who had bad consequences of treating a TB with injections, it might potentially influence their decision." —European and Eastern Mediterranean Regions Participant* |
| **Gender Differences** | Men concerned about modesty of buttocks injection; women more engaged with health system | *"Uh, men with the ladies [females administering injections]. And, of course, at that point, they have lost weight, so their clothes kind of fall off...Uh, they would prefer the arms like other vaccines. That would be very comfortable." —African Region Participant* |
| **LAIs for Children** | Largely favorable despite child aversion to injections; preference over burden of oral dosing | *"But if, um, I do know that my kids might not like injections, but they would get them, um, because it would be easier...I would definitely be interested in injectables because crushing pills is just not great. It's not accurate." —European and Eastern Mediterranean Regions Participant* |
| **Efficacy Equivalence** | Participants require explicit evidence that LAIs are as effective as oral regimens | *"And now I'm transitioning to injection or maybe less pills. What I want to know is how effective that is as well? Is that working? Or was there proof? Uh, were they investigated about the effectiveness? Right? And the other question for me will be, how do I know if the treatment is working besides the controls?" —Americas Region Participant* |
| **Domain: Treatment Characteristic Preferences** |  |  |
| **Treatment Flexibility** | Participants value option to switch between LAI and oral modalities; autonomy over treatment choice is central to acceptability | *"People then, after they are taking the long-acting injectable, don't feel comfortable and want to change the mode of transmission from the injection to taking the pills, or the other way around. Is that possible for people to do that?" —Asia-Pacific Region Participant* |
| **Injection Site Pain** | Pain severity and duration more important than route (IM vs. SC); drug diffusion pain identified as greater barrier than needle penetration | *"It depends on how painful. If it's the pain that is derived from the penetrating the muscle, then that one, they can easily bear because they are used to it…but if it is a pain from the drug getting into the muscles and the pain persists, after the administration, then they might prefer to go back to the pills." —African Region Participant* |
| **Side Effects Impact on Adherence** | Severe side effects after initial injection/series may prevent treatment completion; critical adherence barrier | *"If the side effect is [bad], then people might not have a repeat [come to] a repeat visit if the injection is over a period of time. We have had that problem with TB, with side effects when people can't work properly…so they stop [treatment] immediately because they don't want to be sicker than they are [due to side effects]." —African Region Participant* |
| **Timing & Control of Side Effects** | Patients lose ability to strategically time side effects; LAI schedule is clinic-determined | *"I was on pills. And I could kind of take them at a time, like, right after work, and then I would have the side effects, like, at night. And so I could time that a little bit better...Whereas, you know, if you're on the clinic schedule or whatever and going back to work, you might not have that option." —Americas Region Participant* |
| **Clinic Visit Reduction** | Fewer clinic visits reduce visibility and associated stigma from health workers and community | *"A patient will go to the medical department for injections. How to say — not so often. It means that the risk of receiving stigma and discrimination from medical staff, people, or somebody from his environment [community]. That is why less visits, risk of meeting somebody who will stigmatize him." —European and Eastern Mediterranean Regions Participant* |
| **Privacy & Household Stigma** | LAIs remove visible pill-taking from home/work environments; protects from family/community discrimination | *"It gives them the right to privacy, and getting injections gives them space to do other chores as well as to reduce the cost of frequent visits to the facility. So, most of the women here will prefer injections because they provide that privacy and respect and dignity, and no one can stigmatize them or blame them." —African Region Participant* |
| **Domain: Implementation Considerations** |  |  |
| **Location Preferences** | Mixed preferences depending on health system structure and personal burden; some prefer video DOT with oral regimen to clinic-based LAI | *"That's actually a lot and big, it's kind of a pain. If I could take the pills by myself and on a, you know, DOT, you know, if they can watch me take it, and I can do it from my home, video DOT is way more convenient. For me personally, and I know a lot of working people, having a doctor's appointment every month would be a pain." —European and Eastern Mediterranean Regions Participant* |
| **Options & Information** | Strong desire for choice and access to clear information on trade-offs; concern that providers predetermine treatment pathways | *"One thing, like, I really want for other people who have had TB or have TB is just options. And, um, so, you know, if we're viewing this as another option, I think that's a great way to do it. And, um, the other thing is, of course, you know, please remember children, and please remember people who are pregnant. Um, these are populations that always get left out." —Americas Region Participant* |
| **Political Will & Supply Chain** | Reliable supply chain essential; stockouts would undermine trust and treatment continuity | *"I think in addition to what colleagues have already said, I'm also thinking of consistent supplies. And the, um, they should ensure that people when they start [an LAI], there are no issues of stockouts, uh, no issues of maybe other related, like, maybe injections or, um, supply...So, consistent supply and production should be there. We don't want to have people start and then they stop to start another." —African Region Participant* |

Participants emphasized that they require explicit evidence of LAI efficacy equivalent to oral regimens and value the autonomy to switch between modalities if side effects are intolerable, a concern with serious adherence implications, as a negative experience with an initial injection could prevent treatment completion entirely. The severity and duration of post-injection pain, particularly drug diffusion pain into muscle tissue, were identified as more concerning than needle penetration alone. Beyond clinical considerations, participants highlighted stigma reduction as a key benefit of fewer clinic visits, as well as the privacy afforded by injectable formulations in home and workplace settings. Finally, participants across all regions underscored that successful LAI implementation requires reliable supply chains and political commitment; introducing LAIs without ensuring consistent supply could undermine trust in health systems and negate the benefits these new technologies promise.

## Discussion

In this multi-regional mixed-methods study, stakeholders expressed substantial interest in long-acting injectables for both TB treatment and prevention, although interest depended on regimen and delivery characteristics. Preferences were less consistent when options involved trade-offs related to treatment duration, injection burden, route of administration, and location of care. Qualitative findings further explained these components related to preference and showed that acceptability depended on confidence in efficacy, manageable pain and adverse effects, convenient access, clear information, and preservation of individual choice.

The high level of interest in LAIs observed in our study is consistent with the findings of Vermeulen et al. (20). Among 409 potential TPT recipients in India and South Africa, 75% reported that they would “definitely try” injectable long-acting TPT. Similarly, in our study, 79% preferred a single injection to a one-month oral TPT regimen, while 83% reported probable or definite interest in LAIs for TB treatment. Both studies also identified efficacy and prolonged adverse effects as central concerns. Our qualitative findings extend this evidence by linking these concerns to treatment flexibility, including the option to switch to oral treatment, access to clear information, and patient autonomy. Our study also broadens the evidence base by including stakeholders from 55 countries across all six WHO regions and examining LAIs for both TB prevention and treatment.

Our findings were broadly consistent with a pilot mixed-methods study conducted in the Philippines to inform the development of LAI TB treatment target regimen profiles (19). Bayot et al. found generally favourable attitudes towards LAI treatment, while showing that preferences depended on specific regimen and delivery characteristics. In their discrete choice experiment, one injection per visit significantly increased preference for an LAI regimen among active TB patients, TB survivors, and healthcare providers, whereas three injections per visit reduced preference among active TB patients and survivors. One-time treatment, mild injection-site pain, and delivery at a local health centre were also positively valued, while severe injection-site pain was negatively valued. Bayot et al. further noted that shorter treatment duration was not necessarily prioritised over reducing the number of injections per visit. Although injection burden was operationalised differently in our study, preference increased from 69% to 80% when the number of monthly injections was reduced from four to two. Together, these findings highlight injection burden as an important determinant of LAI acceptability alongside treatment duration, tolerability, and delivery setting.

An important additional finding from our study was the lower interest in LAIs among respondents with lived experience of TB. Although interest remained generally favourable in this group, lived TB experience was associated with approximately half the odds of reporting greater LAI interest after adjustment for setting. The qualitative findings offer a potential explanation: participants described how historical injectable regimens for drug-resistant TB were associated with painful administration and severe adverse effects, including hearing loss. This historical legacy was less prominent in earlier studies, which focused more on current product and delivery characteristics (19,20). Our findings therefore emphasise the need in implementation to distinguish future long-acting formulations clearly from older aminoglycoside-based injectable regimens and to engage directly with communities who have experienced injectable TB treatment.

This study has a number of strengths. It included community members, people affected by TB, civil society representatives, healthcare workers, programme staff, and other stakeholders from all six WHO regions. The sequential explanatory design allowed the survey findings to be interpreted alongside detailed accounts of the experiences, values, and contextual factors underlying those preferences. The study also considered both TB treatment and prevention, directly examined trade-offs among multiple regimen characteristics, and explored less-studied issues including stigma, paediatric administration, and preferences for injectable preventive treatment.

This study has several limitations. Participants were recruited through purposive and snowball sampling, including via TB and civil-society networks, and may have been more knowledgeable about or interested in TB innovation than the wider populations they represent. Nevertheless, these networks enabled us to engage community members and representatives actively involved in TB. Several paired scenarios varied multiple attributes simultaneously, limiting our ability to estimate the independent contribution of each attribute. Future research could consider including larger, more diverse samples, with greater representation of people with lived TB experience, children and adolescents, pregnant people, and participants from underrepresented regions.

Our findings identify several priorities for LAI development. Future regimens should offer a clear advantage over existing oral options by meaningfully reducing treatment duration, pill burden, injection number, or healthcare encounters without compromising efficacy, safety, or treatment flexibility. Product development and introduction should minimize injection pain and prolonged adverse effects, account for local delivery capacity, clearly distinguish new LAIs from historical injectable TB treatments, and preserve meaningful choice between oral and injectable approaches. Continued involvement of people affected by TB throughout target regimen profile development, clinical evaluation, and implementation planning will be essential to ensure that LAIs are not only technically viable, but also trusted, accessible, and patient-centred.

## Data Availability

Summary findings and illustrative quotations are included in the manuscript and supplementary material.

## Acknowledgements

We thank the members of the WHO Long-Acting Injectable Tuberculosis Target Regimen Profile Expert Group for their valuable insights and contributions to the development and interpretation of this study. We also thank Abel Kjaersgaard and Seongmin Cho for helpful discussions. Chaelin Kim and Erin V. McConnell contributed equally to this work and share first authorship.

## Contribution [included in the online submission system; not within the manuscript itself]

C.K. designed and conducted the descriptive and statistical analyses of the survey data, contributed to the design of the qualitative phase based on the quantitative findings, and drafted the manuscript. E.V.M. designed, conducted, and undertook preliminary analyses of Survey B and, with N.S.W., contributed to the design and conduct of the focus group discussions. S.G.S., D.F., M.G., and F.M. conceptualized the study, designed Survey A, contributed to the design of Survey B and the focus group discussions, and acquired funding for Survey B and the qualitative phase. L.A., J.G., E.T., and L.T. contributed to survey design and interpretation of the qualitative findings. G.K. and C.F.M. critically reviewed the methodology, quantitative analyses, and manuscript. N.S.W. designed and conducted the focus group discussions, led the qualitative analysis, and contributed to drafting the qualitative sections of the manuscript. C.K., E.V.M., and N.S.W. jointly integrated findings from the quantitative and qualitative phases. All authors reviewed and approved the final manuscript.

## Disclaimer

The authors alone are responsible for the views expressed in this article, and they do not necessarily represent the decisions, policy, or views of the World Health Organization.

