## Supplemental files for "Stakeholder perspectives and preferences for long-acting injectables for tuberculosis treatment and prevention: a multi-regional mixed-methods study"

### Supplementary material

**Appendix A.** Supplementary methods

##### A-1. Survey instrument development

Two online surveys were designed to collect anonymous, self-reported data on stakeholder preferences and the acceptability of LAIs for tuberculosis prevention and treatment. Survey A (Provider/Policy Survey), led by the World Health Organization (WHO), targeted National TB Programme (NTP) managers, civil society members, and healthcare workers. Survey B (Community/Patient Survey), led by the Treatment Action Group (TAG), targeted community members and people affected by TB.

The surveys were designed and reviewed in English, with Survey B professionally translated into French, Russian, and Spanish to maximize accessibility for global participants. Questions addressed LAI treatment characteristics (e.g., route of administration, injection frequency, treatment duration), trade-off preferences relative to oral regimens, location-of-care preferences, side-effect concerns, and sociodemographic characteristics.

##### A-2. Qualitative data collection

A semi-structured FGD facilitation guide was developed to explore Phase 1 survey findings in greater depth, with topic areas including: preferences for LAI treatment characteristics (injection frequency, route of administration, treatment duration), trade-offs between LAIs and oral regimens, location of administration preferences, side effect concerns, stigma-related considerations, and acceptability of LAIs for children. Each focus group comprised two to five participants; facilitators probed for regional variation and encouraged participants to elaborate on the contextual factors shaping their perspectives. All sessions were recorded with participant consent and transcribed. Focus group discussions (FGDs) were conducted via Zoom, which has been shown to be an effective way to conduct qualitative research across diverse stakeholder groups (Reñosa et al., 2021).

##### A-3. Qualitative analytic rigor and reflexivity

To enhance rigor and transparency, the coding process was documented through analytic memos written by N.S.W. during and after initial familiarization with data, which tracked emerging patterns, analytical decisions, and areas of interpretive disagreement during codebook development through discussions with E.V.M. Memos from data familiarization were used to refine code definitions. The analysis team spans research, programmatic, and activism experience in implementing TB innovations, which informed both the a priori domains and the interpretation of findings. Our existing knowledge and study objectives shaped our view of the data and our interpretations. As the primary coder, N.S.W. maintained a reflexive journal throughout the coding process, documenting positionality, emerging insights, and tensions encountered. The journal captured instances where code definitions required refinement, potential conflicting interpretations of participant statements, and reflections on positionality.

**Appendix B.** Factors Associated with Interest in LAI for TB Treatment (Ordinal Logistic Regression)

**B-1. Model Selection Strategy Comparison**

Stepwise AIC-based selection using backward, forward, and bidirectional approaches identified the same final model, retaining setting and TB experience as predictors of interest.

| Model Strategy | Variables Included | AIC | N Parameters |
|---|---|---|---|
| 1. p<0.05 only | region, tb_experience | 501.48 | 5 |
| 2. p<0.20 only | region, tb_experience, setting | 500.91 | 7 |
| 3. Full model | region, age_range, gender, tb_experience, setting | 505.30 | 10 |
| 4. Backward | tb_experience, setting | 498.11 | 3 |
| 5. Forward | tb_experience, setting | 498.11 | 3 |
| 6. Both directions | tb_experience, setting | 498.11 | 3 |
| 7. A priori | region, age_range, gender | 507.57 | 7 |

**B-1.1. Crude Model P-values:**

- Age Range: p = 0.325
- Gender: p = 0.894
- Setting: p = 0.053
- TB Experience: p = 0.047 *
- WHO Region: p = 0.034 *

**B-1.2. Conclusion:**

Best model by AIC: 4. Backward (AIC = 498.11)
Selected variables: tb_experience, setting

**B- 1.3. Strategy:**

Strategy 1: Include only variables with p<0.05 in crude analysis
Strategy 2: Include variables with p<0.20 in crude analysis
Strategy 3: Include all variables regardless of crude p-values
Strategy 4-6: Stepwise selection based on AIC
Strategy 7: Include theoretically important demographic variables

* indicates p < 0.05

**Appendix C.** Characteristics of survey and focus group participants

##### C-1 Table. Survey participant characteristics by primary role or affiliation and WHO region

| Characteristic | Survey A — Provider/Policy (WHO-led), N = 102 n (%) | Survey B — Community/Patient (TAG-led), N = 208 n (%) | Total N = 310 n (%) |
|---|---|---|---|
| **Primary role / affiliation** | | | |
| Civil society | 26 (25%) | 95 (46%) | 121 (39%) |
| Clinician / healthcare worker | 26 (25%) | 50 (24%) | 76 (25%) |
| Person with lived TB experience | 0 (0%) | 60 (29%) | 60 (19%) |
| Multilateral organisation or NGO | 18 (18%) | 0 (0%) | 18 (6%) |
| NTP staff | 16 (16%) | 0 (0%) | 16 (5%) |
| Laboratory | 11 (11%) | 0 (0%) | 11 (4%) |
| Researcher | 5 (5%) | 3 (1%) | 8 (3%) |
| **WHO region** | | | |
| African Region | 11 (11%) | 91 (44%) | 102 (33%) |
| European Region | 27 (26%) | 49 (24%) | 76 (25%) |
| South-East Asia Region | 17 (17%) | 30 (14%) | 47 (15%) |
| Western Pacific Region | 34 (33%) | 6 (3%) | 40 (13%) |
| Region of the Americas | 3 (3%) | 30 (14%) | 33 (11%) |
| Eastern Mediterranean Region | 0 (0%) | 2 (1%) | 2 (1%) |
| Global / multi-region | 10 (10%) | 0 (0%) | 10 (3%) |
| **Total** | **102 (100%)** | **208 (100%)** | **310 (100%)** |

##### C-2 Table. Additional characteristics of community/patient survey participants

| Characteristic | n (%) |
|---|---|
| **Gender** | |
| Female | 107 (51%) |
| Male | 98 (47%) |
| Gender minority (transgender / non-binary) | 3 (1%) |
| **Age group (years)** | |
| 18–25 | 15 (7%) |
| 26–34 | 46 (22%) |

| | |
|---|---|
| 35–44 | 59 (28%) |
| 45–54 | 57 (27%) |
| 55–64 | 23 (11%) |
| 65+ | 8 (4%) |
| **Setting (analytic n = 205)** | |
| Urban | 133 (65%) |
| Rural | 43 (21%) |
| Other | 29 (14%) |
| **Prior awareness of LAIs for TB** | |
| Aware | 110 (53%) |
| Not aware | 98 (47%) |
| **Prior TB treatment or TB preventive treatment** | |
| Yes | 79 (38%) |
| No | 129 (62%) |
| **Prior experience with injectables for TB** | |
| Yes | 21 (10%) |
| No | 187 (90%) |

**C-3 Table. Qualitative focus group participant characteristics**

| Focus group (WHO regional grouping) | Participants, n | Gender | Median age range (years) |
|---|---|---|---|
| African Region (AFRO) | 4 | Female 3 / Male 1 | 55–64 |
| Western Pacific & South-East Asia Regions (WPRO/SEARO) | 5 | Female 4 / Male 1 | 35–44 |
| Region of the Americas (AMRO) | 3 | Female 2 / Male 1 | 35–44 |
| European & Eastern Mediterranean Regions (EURO/EMRO) | 2 | Female 2 / Male 0 | 45–54 |
| **Total** | **14** | **Female 11 (79%) / Male 3 (21%)** | **35–44** |

*No gender-minority participants took part in the focus groups.*

**Notes:** Percentages may not sum to 100% due to rounding. LAI = long-acting injectable; TB = tuberculosis; NTP = National TB Programme; NGO = non-governmental organisation; FGD = focus group discussion.

Survey A (Provider/Policy) was led by WHO and targeted NTP managers, civil society members and healthcare workers; Survey B (Community/Patient) was led by the Treatment Action Group (TAG) and recruited community members and persons affected by TB. For questions with identical wording and response options, the two surveys were combined (N = 310). Focus group participants were recruited from Survey B respondents who indicated willingness to be re-contacted and are therefore a subset of, not additional to, the survey sample. Setting was available for 205 of 208 community respondents (3 missing).

**Appendix D. Interest in LAI for TB Treatment by Stakeholder Characteristics**

| Characteristics | Definitely not n (%) | Probably not n (%) | I am unsure n (%) | Probably n (%) | Definitely n (%) | Mean (SD) | n |
|---|---|---|---|---|---|---|---|
| Overall | 8 (2.6) | 17 (5.5) | 29 (9.4) | 119 (38.4) | 137 (44.2) | 4.16 (0.98) | 310 |
| **Primary Role** | | | | | | | |
| Researcher | 0 (0.0) | 1 (12.5) | 1 (12.5) | 4 (50.0) | 2 (25.0) | 3.88 (0.99) | 8 |
| Person with TB experience | 4 (6.7) | 4 (6.7) | 7 (11.7) | 21 (35.0) | 24 (40.0) | 3.95 (1.19) | 60 |
| NTP staff | 0 (0.0) | 1 (6.2) | 0 (0.0) | 11 (68.8) | 4 (25.0) | 4.12 (0.72) | 16 |
| Multilateral organisation or NGO | 0 (0.0) | 1 (5.6) | 0 (0.0) | 13 (72.2) | 4 (22.2) | 4.11 (0.68) | 18 |
| Laboratory | 0 (0.0) | 0 (0.0) | 3 (27.3) | 5 (45.5) | 3 (27.3) | 4.00 (0.77) | 11 |
| Clinician/health worker | 1 (1.3) | 5 (6.6) | 7 (9.2) | 25 (32.9) | 38 (50.0) | 4.24 (0.96) | 76 |
| Civil society | 3 (2.5) | 5 (4.1) | 11 (9.1) | 40 (33.1) | 62 (51.2) | 4.26 (0.96) | 121 |
| **WHO Region** | | | | | | | |
| Africa | 4 (3.9) | 6 (5.9) | 7 (6.9) | 33 (32.4) | 52 (51.0) | 4.21 (1.07) | 102 |

| | | | | | | | |
|---|---|---|---|---|---|---|---|
| Americas | 0 (0.0) | 3 (9.1) | 6 (18.2) | 13 (39.4) | 11 (33.3) | 3.97 (0.95) | 33 |
| Eastern Mediterranean | 0 (0.0) | 0 (0.0) | 1 (50.0) | 0 (0.0) | 1 (50.0) | 4.00 (1.41) | 2 |
| Europe | 1 (1.3) | 2 (2.6) | 10 (13.2) | 30 (39.5) | 33 (43.4) | 4.21 (0.87) | 76 |
| Global | 0 (0.0) | 2 (20.0) | 0 (0.0) | 6 (60.0) | 2 (20.0) | 3.80 (1.03) | 10 |
| South-East Asia | 3 (6.4) | 1 (2.1) | 3 (6.4) | 16 (34.0) | 24 (51.1) | 4.21 (1.10) | 47 |
| Western Pacific | 0 (0.0) | 3 (7.5) | 2 (5.0) | 21 (52.5) | 14 (35.0) | 4.15 (0.83) | 40 |

*Note: n = frequency; Percentages may not sum to 100% due to rounding. LAI = Long-acting injectable; TB = Tuberculosis*

**Appendix E. Factors Associated with Interest in LAI for TB Treatment (Community Survey)**

| | | | Univariate | | Multivariate* | |
|---|---|---|---|---|---|---|
| **Variable** | **n (%)** | **Mean (SD)** | **COR (95% CI)** | **P-value** | **AOR (95% CI)** | **P-value** |
| **Age Range** | | | | | | |
| 18-34 | 60 (29.3%) | 4.20 (1.01) | 1.00 (reference) | | - | |
| 35-54 | 114 (55.6%) | 4.22 (1.03) | 1.08 (0.60-1.96) | 0.79 | - | - |
| 55+ | 31 (15.1%) | 4.06 (0.89) | 0.67 (0.30-1.48) | 0.32 | - | - |
| **Gender** | | | | | | |
| Female | 107 (52.2%) | 4.20 (0.97) | 1.00 (reference) | | - | |
| Male | 98 (47.8%) | 4.18 (1.04) | 1.04 (0.62-1.74) | 0.89 | - | - |
| **Setting** | | | | | | |
| Urban | 133 (64.9%) | 4.22 (0.99) | 1.00 (reference) | | 1.00 (reference) | |
| Other | 29 (14.1%) | 3.86 (1.06) | 0.49 (0.24-1.01) | 0.05 | 0.46 (0.22-0.95) | 0.04 |
| Rural | 43 (21.0%) | 4.33 (0.97) | 1.25 (0.65-2.43) | 0.51 | 1.34 (0.70-2.64) | 0.39 |
| **TB Experience** | | | | | | |
| Person without reported lived experience with TB | 146 (71.2%) | 4.29 (0.90) | 1.00 (reference) | | 1.00 (reference) | |
| Person with lived experience with TB | 59 (28.8%) | 3.93 (1.19) | 0.56 (0.32-0.99) | 0.05 | 0.51 (0.29-0.91) | 0.02 |
| **WHO Region** | | | | | | |
| Africa | 91 (44.4%) | 4.16 (1.08) | 1.00 (reference) | | - | |
| Americas | 28 (13.7%) | 3.82 (0.94) | 0.44 (0.20-0.94) | 0.03 | - | - |
| Europe | 49 (23.9%) | 4.39 (0.73) | 1.29 (0.67-2.50) | 0.46 | - | - |
| Other | 7 (3.4%) | 4.43 (0.79) | 1.46 (0.34-7.53) | 0.62 | - | - |
| South-East Asia | 30 (14.6%) | 4.23 (1.17) | 1.31 (0.59-3.01) | 0.51 | - | - |

*Multivariate model includes variables selected by stepwise AIC selection: Setting and TB Experience. Variables not selected (Age Range, Gender, WHO Region) show '-' in multivariate columns.

Abbreviations: COR, Crude Odds Ratio; AOR, Adjusted Odds Ratio; CI, Confidence Interval. Interest score based on 5-point scale (Definitely not to Definitely). Odds ratios >1 indicate higher odds of greater interest in LAI treatment. Reference categories: 18-34 years (Age Range), Female (Gender), Urban (Setting), Person without reported lived experience with TB (TB Experience), Africa (WHO Region). n = 205. Model AIC = 498.11.

**Appendix F. Treatment preference questions as presented to respondents**

Survey on Community Perceptions and Preferences Regarding Long-Acting Technologies for TB Treatment and Prevention

**Preferences on Long-Acting Injectables (LAIs) in TB Treatment**

*We want to understand your preferences for LAIs compared to the current standard of care. The following questions will ask about your preferences regarding LAIs.*

*For each question, we will provide you with two options (options A and B) and ask you to respond using a five-point preference scale (strongly prefer option A, Prefer option A, Neutral, Prefer option B, strongly prefer option B).*

***For this part, please assume that all the injectable treatments described work just as well and are just as safe as the TB pills someone might take now.***

1) **Standard of care vs LAI regimen without co-formulation**

We want to know which you would prefer: the current 6-month pill treatment or a new possible 3-month treatment with injections. At first, each medicine may need to be given as a separate shot to work as well as your daily pills.

Under this scenario, please consider the following two options:

**Option A.** Standard of care treatment. The oral treatment duration for TB is 6 months. You need to take medication every day.

**Option B.** A three-month treatment regimen in which you would take four medicines orally (by mouth) every day for the first month, followed by four injections in the second month, and the same four injections in the third month.

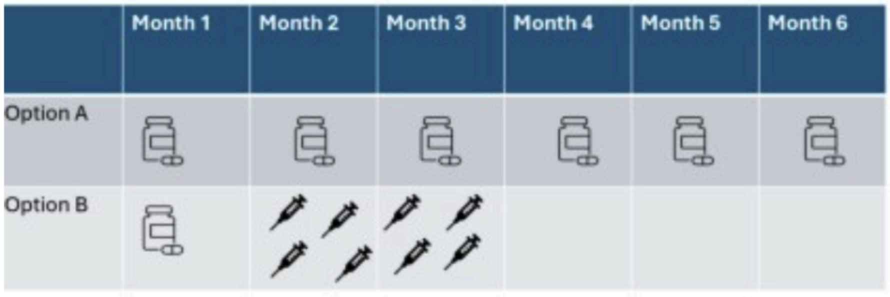

2) **Standard of care vs LAI regimen with co-formulation**

In the future, it may be possible to have fewer injections that each include multiple medications, meaning fewer injections needed each month.

Under this scenario, please consider the following two options:

**Option A.** Standard of care treatment. The oral treatment duration for TB is 6 months. You need to take medication by mouth every day for 6 months (e.g., a pill or more than one pill).

**Option B.** A three-month treatment regimen in which you would take medicines orally (by mouth) daily for the first month, followed by two injections in the second month, and the same two injections in the third month.

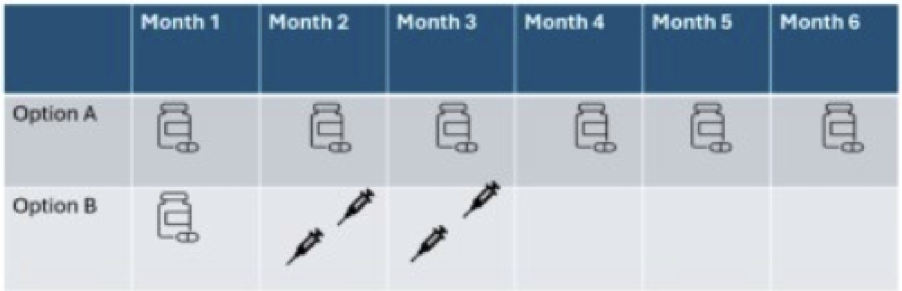


3) **Coverage duration vs number of injections**

During the development of these treatments, researchers may learn that LAIs require more injections in a single clinical visit for longer protection - meaning, it could be six injections in a single visit for two months of treatment coverage instead of three injections. This question asks you to evaluate the trade-off between the time the treatment lasts and the number of injections required. Some medications may need two injections simultaneously to extend the effectiveness of the treatment from one month to two months.

In this context, please consider the following two options:

**Option A:** A four-month treatment regimen that includes three injections each month.

**Option B:** A three-month treatment regimen that includes six injections administered over two clinic visits, two months apart.

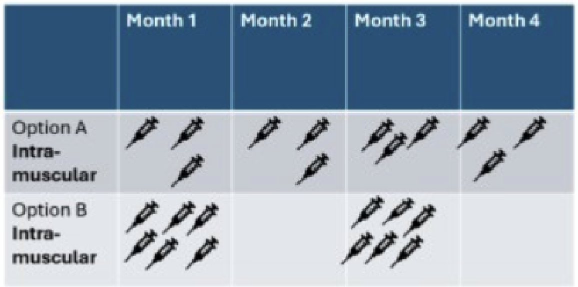


4) **Route of administration**

**Subcutaneous injections**, shallow injections just under your skin using shorter needles. These often feel less painful.

**Intramuscular injections,** deeper injections into your muscle using longer needles. These may allow the medicine to work longer in your body.

In this context, please consider the following two options:

**Option A:** A four-month treatment regimen consisting of three **subcutaneous** injections given once a month over four months.

**Option B.** A three-month treatment regimen that includes three injections administered **intramuscular** in the first month and repeated two months later.

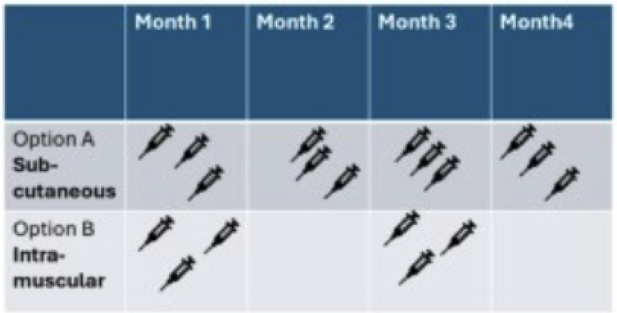


5) **Health Facility: community vs. centralized**

It may be determined that drugs with longer time between injections, such as going two months compared to one month between injections may mean that treatment needs to be delivered at **a hospital**.

If you could go to the hospital two times in four months for treatment as compared to going to the local clinic once a month, each month, for four months, please consider the two options:

**Option A**. A four-month regimen includes three injections per visit administered at the ***local clinic*** once a month for four months.

**Option B.** A three-month regimen that includes three injections per visit at the ***hospital*** done two times, two months apart (month 1 and month 3).

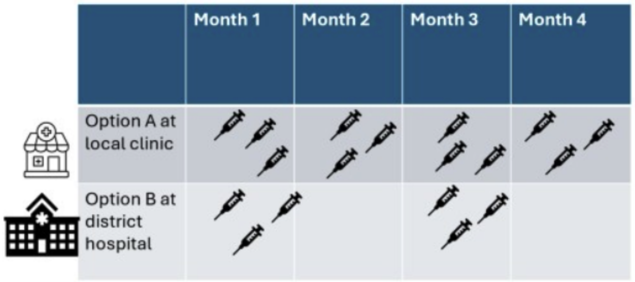


6) **Single injection visit for the continuation phase**

When starting a long-acting injectable (LAI) treatment, you might begin with taking pills by mouth (the "intensive, oral lead-in phase"). After this pill-taking period, you might only need one injection visit to complete your treatment (the "continuation phase").

This approach could mean fewer injections overall - just a single injection visit at the end of your treatment after taking pills for a while, rather than multiple injection appointments throughout your treatment.

In this context, please consider the following two options:

**Option A:** A three-month regimen in which you would take medicines orally (by mouth) daily for the first month, followed by three injections, administered monthly for two months.

**Option B:** A three-month regimen in which you would take medicines orally (by mouth) daily for two months, followed by a single visit during which three injections are administered.

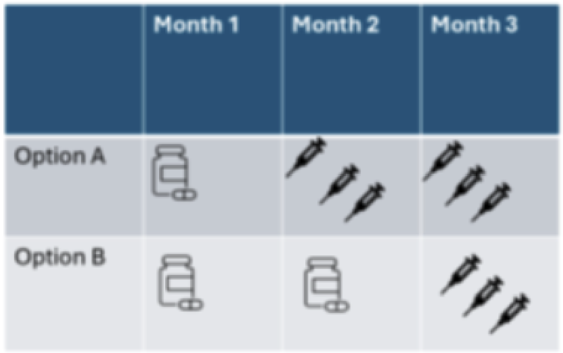

7) **Long-Acting Injectables (LAIs) for prevention of TB disease**

It may be determined that an LAI could be used for TB preventive treatment (TPT).

Please indicate your preference.

**Option A:** A one-month all-oral regimen for prevention of TB

**Option B:** A single LAI injection for prevention of TB
